# Introduction into the Marseille geographical area of a mild SARS-CoV-2 variant originating from sub-Saharan Africa

**DOI:** 10.1101/2020.12.23.20248758

**Authors:** Philippe Colson, Anthony Levasseur, Philippe Gautret, Florence Fenollar, Van Thuan Hoang, Jeremy Delerce, Idir Bitam, Rachid Saile, Mossaab Maaloum, Abdou Padane, Marielle Bedotto, Ludivine Brechard, Vincent Bossi, Mariem Ben Khedher, Hervé Chaudet, Matthieu Million, Hervé Tissot-Dupont, Jean-Christophe Lagier, Souleymane Mboup, Pierre-Edouard Fournier, Didier Raoult

## Abstract

**BACKGROUND:** In Marseille, France, the COVID-19 incidence evolved unusually with several successive epidemic episodes. The second outbreak started in July, was associated with North Africa, and involved travelers and an outbreak on passenger ships. This suggested the involvement of a new viral variant.

**METHODS:** We sequenced the genomes from 916 SARS-CoV-2 strains from COVID-19 patients in our institute. The patients’ demographic and clinical features were compared according to the infecting viral variant.

**RESULTS:** From June 26^th^ to August 14^th^, we identified a new viral variant (Marseille-1). Based on genome sequences (n=89) or specific qPCR (n=53), 142 patients infected with this variant were detected. It is characterized by a combination of 10 mutations located in the *nsp2, nsp3, nsp12, S, ORF3a, ORF8* and *N/ORF14* genes. We identified Senegal and Gambia, where the virus had been transferred from China and Europe in February-April as the sources of the Marseille-1 variant, which then most likely reached Marseille through Maghreb when French borders reopened. In France, this variant apparently remained almost limited to Marseille. In addition, it was significantly associated with a milder disease compared to clade 20A ancestor strains.

**CONCLUSION:** Our results demonstrate that SARS-CoV-2 can genetically diversify rapidly, its variants can diffuse internationally and cause successive outbreaks.

## 1. Introduction

The SARS-CoV-2 virus emerged in humans in Wuhan, China, in December 2019, prior to speading worldwide. In China and Europe, the epidemic had a bell shape typical of a respiratory virus (https://covid19-country-overviews.ecdc.europa.eu/) [1, 2]. Surprisingly, in the countries that closed their borders, the evolution varied: in some countries, no other epidemic was detected whereas in others, new epidemic waves occurred, caused by new variants [3]. Among the new sources that may explain the occurrence of different epidemics according to geographical zones, the role of intensive animal breeding like mink farming in Denmark [4] remains a mystery. In Marseille, the bell-shaped curve ended in May, but new cases and then an atypically-shaped epidemic reappeared upon the border reopening. The reopening of borders with Maghreb occurred despite the fact that a very active COVID-19 outbreak was ongoing in Algeria. Interestingly, the first cases of the July epidemic had direct or indirect contacts with passengers from ferries coming from Tunisia or Algeria, which led us to suspect that this variant had an African origin. In our institute (Méditerranée Infection Institute [IHU]) in Marseille, Southern France, we investigated the viral genotypes from patients diagnosed in Marseille using genomic sequencing and genotype-specific PCR. Then, we also tested patients’ specimens from Algerian, Moroccan and Senegalese residents for the presence of a new variant that we named Marseille-1.

## 2. Materials and methods

### 2.1 Virological diagnosis of SARS-CoV-2 infections

In IHU in Marseille, France, we have carried out SARS-CoV-2 RNA testing using real-time reverse transcription-PCR (qPCR) since the end of January 2020, as previously described [1, 5]. The numbers of tests and cases were daily monitored since the first positive diagnosis on 02/27/2020 [2] (https://www.mediterranee-infection.com/covid-19/).

### 2.2 Study period and clinical samples

Whole genome sequencing of SARS-CoV-2 genome was performed from nasopharyngeal samples tested between June and August 2020 at IHU. Specimens with a cycle threshold value (Ct) lower than 20 were selected in priority, and those with a Ct between 20 and 30 were included secondarily to cover the study period more comprehensively. The study was approved by the ethical committee of the University Hospital Institute Méditerranée Infection (N°: 2020-016-2).

### 2.3 Genome sequencing and analysis

Viral genomes were obtained using next-generation sequencing (NGS) and the Illumina technology (Illumina Inc., San Diego, CA, USA), as previously described [2, 6]. Viral RNA was extracted from 200 µL of nasopharyngeal swab fluid using the EZ1 Virus Mini Kit v2.0, and was reverse transcribed using SuperScript IV (ThermoFisher Scientific, Waltham, MA, USA) prior to cDNA second strand synthesis with Klenow Fragment DNA polymerase (New England Biolabs, Beverly, MA, USA). The generated DNA was purified using Agencourt AMPure XP beads (Beckman Coulter, Villepinte, France) and sequenced using the Illumina Nextera XT Paired end strategy on a MiSeq instrument. Genome consensus sequences were generated with the CLC Genomics workbench v.7 by mapping on the SARS-CoV-2 genome GenBank accession no. NC_045512.2 (Wuhan-Hu-1 isolate) with the following thresholds: 0.8 for coverage and 0.9 for similarity. SARS-CoV-2 sequences obtained in our institute have been submitted to the GISAID database.

Sequences from complete genomes were analyzed using the Nextclade web-tool (https://clades.nextstrain.org/) [7]. Clades were defined based on the occurrence of at least five genomes sharing the same pattern of mutations. These genome sequences were compared to those available in the GISAID database (https://www.gisaid.org/). Phylogenetic trees were reconstructed by using Nextclade and visualized with iTOL (https://itol.embl.de/).

### 2.4 Marseille-1 variant specific RT-PCR

For the specimens with Ct values > 30 or those with Ct values < 30 but from which genome sequences were not obtained, we attempted to identify those harboring the Marseille-1 variant using RT-PCR targeting a fragment of the nucleocapsid-encoding gene harboring two mutations separated by 17 nucleotides concurrently present in the Marseille-1 variant. RT-PCR was performed with the PrimerF1 (forward, 5’-TCTACGCAGAAGGGAGCAGA-3’) and PrimerR1 (reverse, 5’-GGAGAAGTTCCCCTACTGCTG-3’) primers, and the QuantiNova SYBR Green RT-PCR kit (Qiagen, Hilden, Germany). In order to evaluate whether the Marseille-1 variant was also prevalent in these countries, this PCR system was also applied to 97 SARS-Cov-2 specimens from COVID-19-positive residents from Senegal, 278 from Algeria and 94 from Morocco. All specimens had been sampled in October and November 2020.

### 2.5 Comparisons of epidemiological and clinical features of patients diagnosed during episodes 1 and 2

From February 29^th^ to August 31^st^, 2020, the demographic and clinical features of the patients infected with the Marseille-1 variant were compared to those of the patients infected during episode 1 with 20A variants.

### 2.6 Statistical analyses

Statistical tests were done using R 4.0.2 (https://cran.r-project.org/bin/windows/base/): Chi2 or Fisher’s exact test for qualitative variables, and Student’s t-test for quantitative variables. A p<0.05 was considered statistically significant.

### 2.7 Ethics statement

The study was approved by the ethical committee of the Méditerranée Infection Institute (N°: 2020-016-2). Access to the patients’ biological and registry data issued from the hospital information system was approved by the data protection committee of Assistance Publique-Hôpitaux de Marseille (APHM) and was recorded in the European General Data Protection Regulation registry under number RGPD/APHM 2019-73.

## 3. Results

### 3.1 Epidemiological evidence of an African source

The Marseille-1 variant emerged in week 27, when it accounted for 100% of sequenced genomes (**Figure 1**). Between June 24^th^ and July 15^th^, information on recent travel abroad by SARS-CoV-2-positive patients was documented at our institute for 939 of 1,108 cases. This period coincided with an increase in the number of positive SARS-CoV-2 tests from patients returning from Tunisia, with 14 cases, following the reopening of borders, and the resumption of passenger boat traffic between France and this country (**Supplementary Figure 1, Supplementary Table 1**). Other patients who traveled by ferry between North Africa (Tunisia and Algeria) and Marseille were diagnosed between July 8^th^ and 20^th^. In addition, two patients diagnosed on July 3^rd^ and July 7^th^, who had no travel history, reported a contact with SARS-CoV-2-positive Algerian travelers as source of their COVID-19. Another 35 positive patients worked or traveled on ships from the same passenger ferry company that sailed between Maghreb and Marseille. Such an evidence of an African origin for the Marseille-1 variant motivated a genotypic study to evaluate the incidence and history of this variant.

**Table 1.**
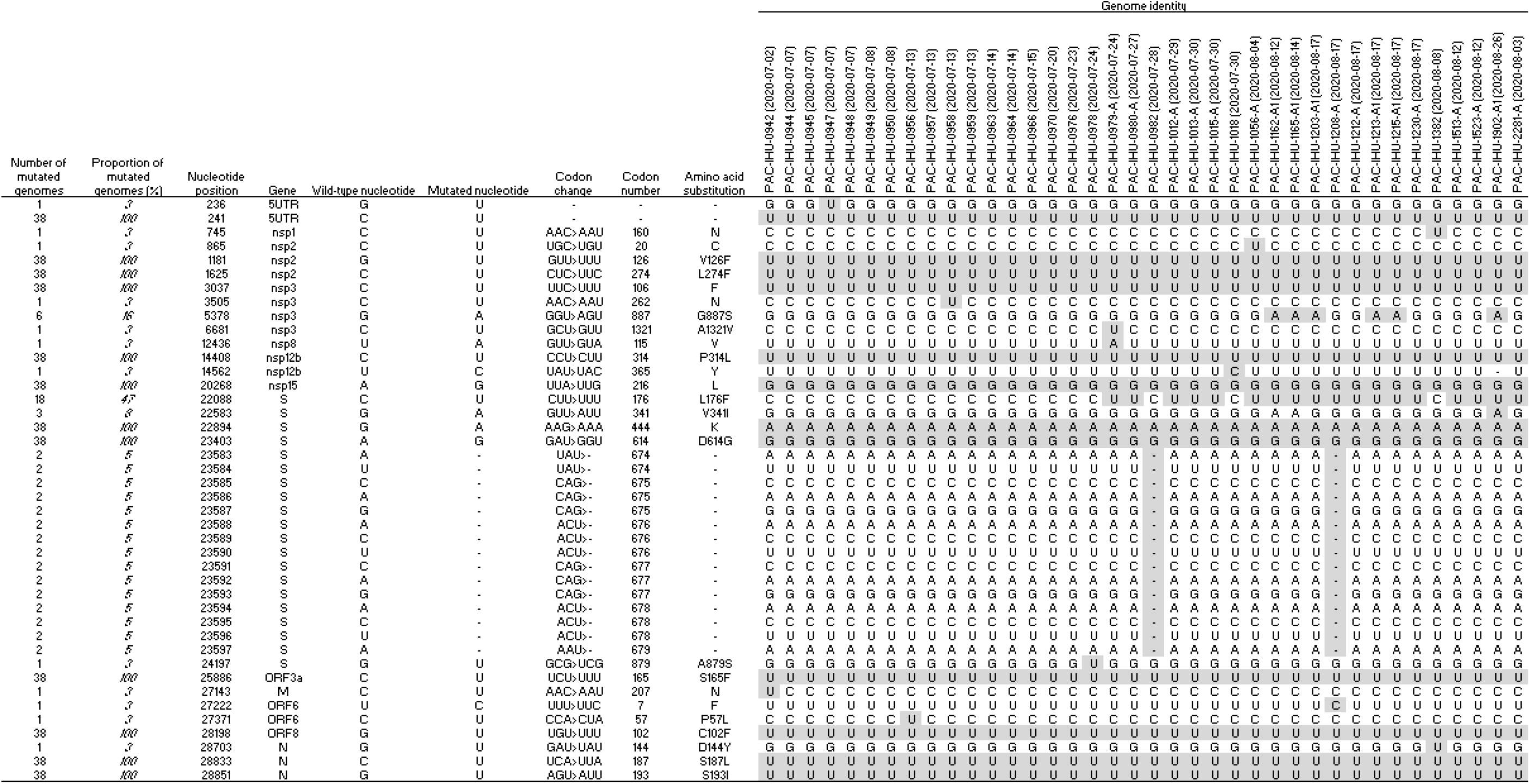
Nucleotide mutations and amino acid substitutions in the genomes of Marseille-1 variants

**Figure 1.**
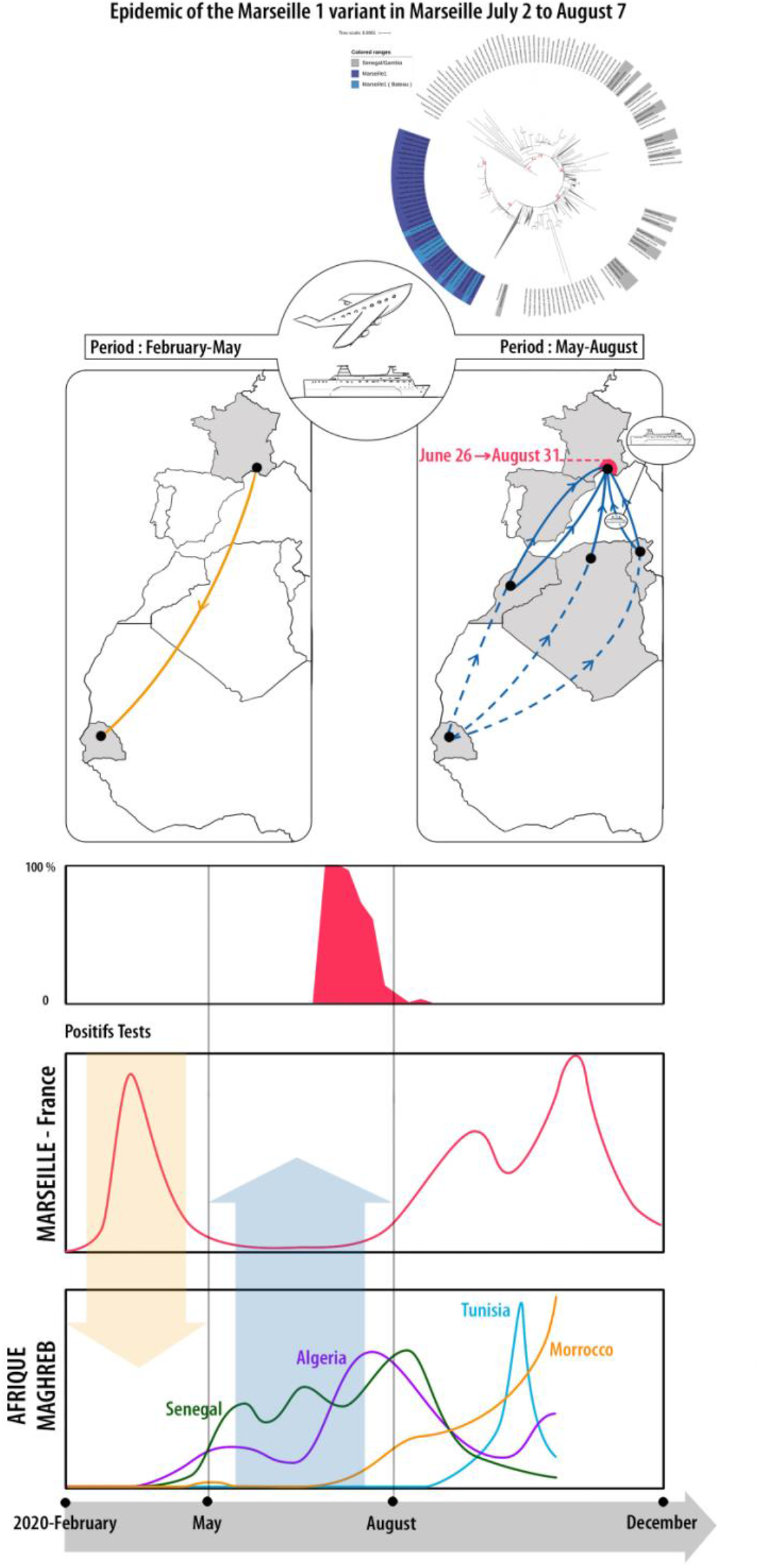
The SARS-CoV-2 Marseille-1 variant outbreak. A. Left: transfer of the 20A variant from France to Senegal and Gambia during episode 1; right: proposed transfer route of the Marseille-1 variant from sub-Saharan Africa to Maghreb (dashed lines), and then to France. B. Evolution of the proportion of Marseille-1 variant genomes identified in Marseille over time. The time scale is featured at the bottom of the figure. C. Epidemic curve of the COVID-19 in Marseille, France. D. Epidemic curves of the COVID-19 in Senegal, Morocco, Algeria and Senegal. The yellow arrow shows the transfer of the 20A variant from France to Senegal whereas the blue arrow shows the transfer of the Marseille-1 variant to France.

### 3.2 Outcome of the Marseille-1 variant

As of December 17^th^, 2020, 405,070 tests were performed for SARS-CoV-2 infection for 289,689 patients, of whom 25,446 (8.8%) were positive (**Figure 2**). We have diagnosed 6,855 patients during episode 1 and 18,591 during episode 2. We obtained SARS-CoV-2 full-length genome sequences from 916 patients (submitted to the GISAID database). These included 429 genomes from episode 2, which were added to 487 genomes from episode 1 ^11^. Time-scaled phylogeny enabled differentiating ten clusters, identified as variants Marseille-1 to Marseille-10, encompassing at least 5 genomes each [3]. Between July and August, the Marseille-1 variant predominated, with 38 complete genome sequences being obtained. In addition, 51 draft genomes were identified as of the Marseille-1 variant, and this genotype was identified from an additional 53 samples by specific RT-PCR. Overall, 142 patients were detected as infected with the Marseille-1 variant. In addition, seven specimens/97 from Senegal, 44/278 from Algeria, and 15/94 from Morocco, all sampled in October and November 2020, were also RT-PCR-positive for the Marseille-1 variant (**Supplementary Figure 1**).

**Figure 2.**
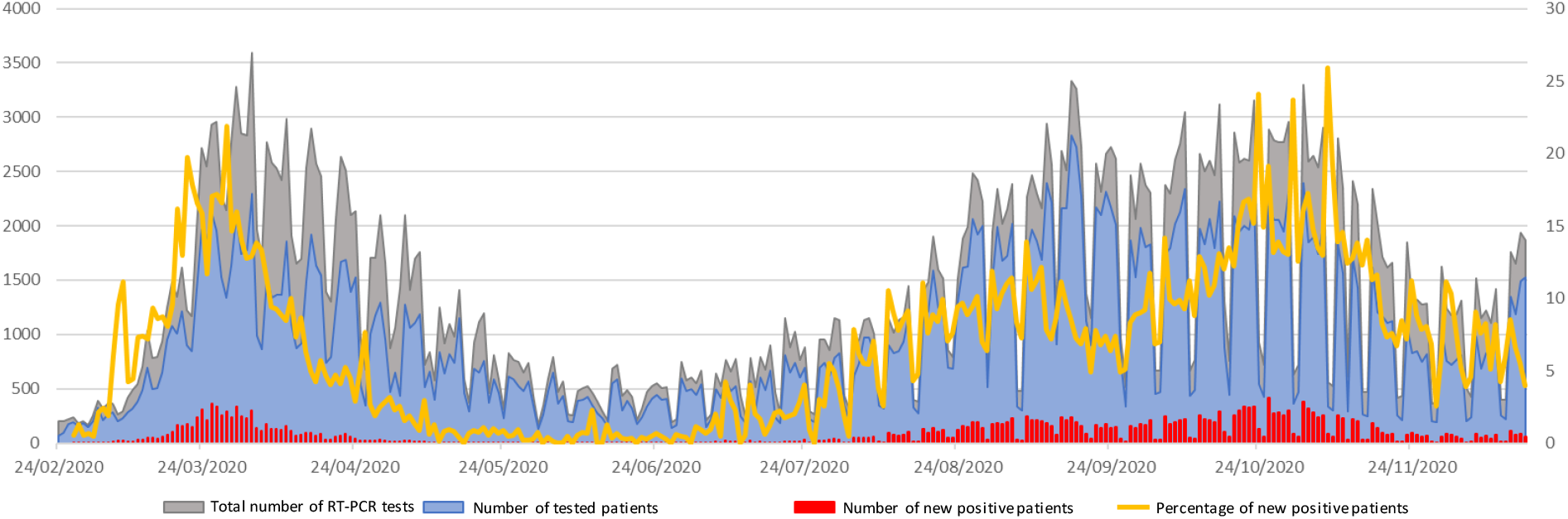
**Number of RT-PCR tests, tested patients, positive patients and percentage of positive patients from February 24^th^ to December 17^th^, 2020**

### 3.3 Phylogenetic study of the Marseille-1 variant

Phylogeny recontruction using genomic sequences available in GISAID showed that Marseille-1 variants belonged to a cluster that comprised almost only sequences from sub-Saharan Africa including from Senegal and Gambia as well as from the Marseille area (**Figure 3, Supplementary Figure 2**). However, RT-PCR results demonstrated that Marseille-1 variants were also detected in travelers from Tunisia to Marseille as well as in Algerian and Moroccan residents. According to the date when sequenced strains were sampled, the earliest ancestors of Marseille-1 strains detected in the GISAID database were from samples collected in Senegal on March 31^st^, 2020, and in Gambia on June 1^st^, 2020 (**Figure 1**).

**Figure 3.**
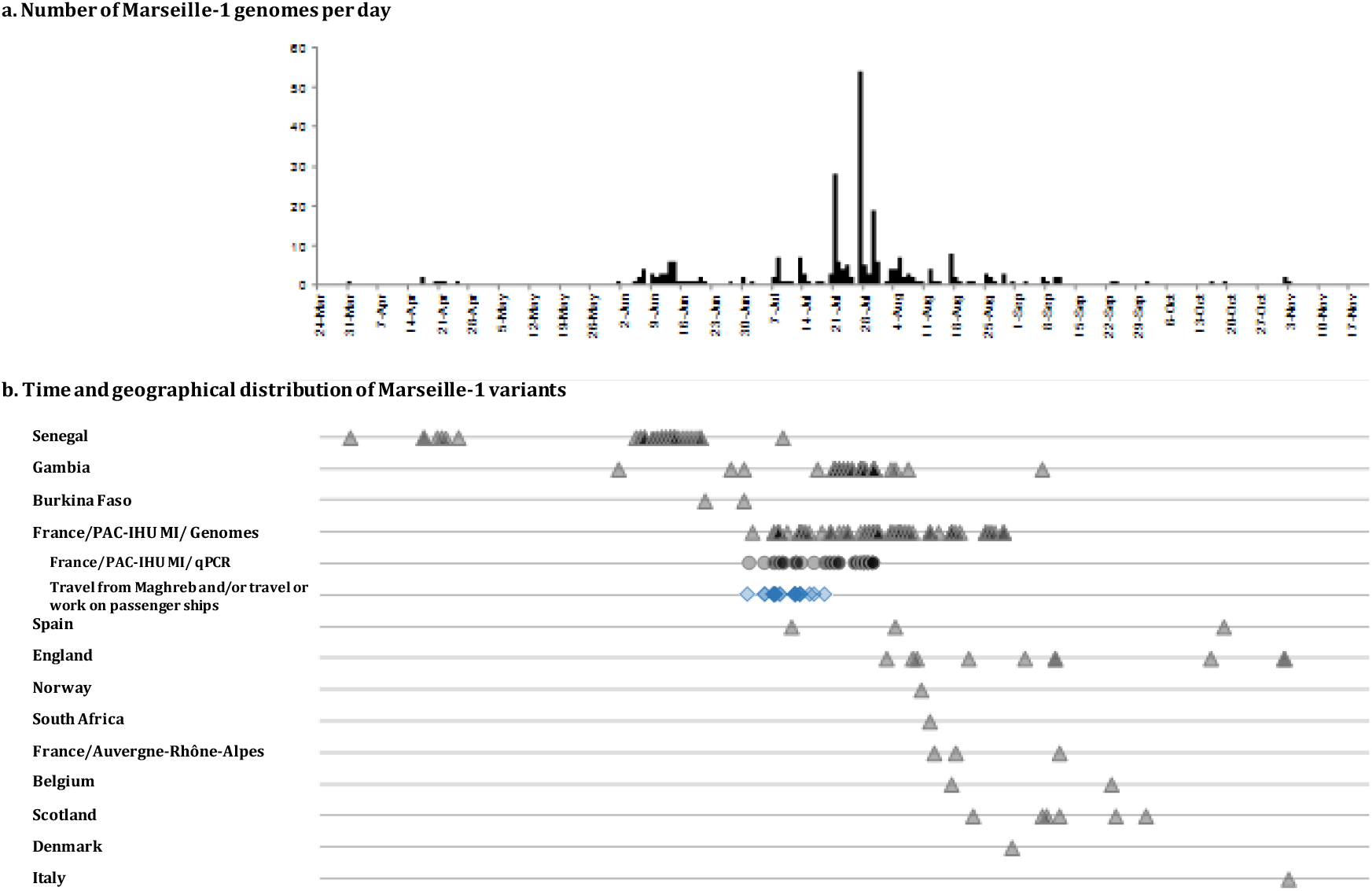
Evolution of the Marseille-1 variant epidemic, from Sub-Saharan Africa to Marseille. a) Numbers of patients with identified Marseille-1 strains detected over time in our institute; b) numbers of Marseille-1 genomes detected in our institute and in the GISAID database according to time of deposit and country. Triangles and circles indicate Marseille-1 strains identified using genome sequences or variant-specific PCR, respectively. Travel from Maghreb and/or travel or work on passenger ships is indicated by blue diamonds.

Several studies have described that clade 20A strains were brought to sub-Saharan Africa, particularly Senegal and Gambia, mostly by French travelers during episode 1 **[8],[9],[10]**. We assume that the ancestors of Marseille-1 variants originated in Senegal or Gambia, through progressive accumulation of mutations by clade 20A strains, and that their offspring were then brought to Maghreb and imported to Marseille by travelers as well as ferry sailors from Algeria and Tunisia (**Figure 1**). Such an hypothesis is supported by several arguments. First, a recent study of SARS-CoV-2 genomes suggested that sub-Saharan viral strains may have followed human routes through Mali, Tunisia or Egypt to Europe **[10]**. In addition, the emergence of Marseille-1 variants in Marseille followed the resumption in week 27 of maritime and air connections between Maghreb and Marseille (Supplementary Table 1). However, despite the diffusion of the Marseille-1 epidemic in the population from Marseille, it seemingly barely spread outside the city, its prevalence rapidly decreased, being <10% for week 31 and it disappeared at the end of August.

### 3.4 Genotypic and amino acid patterns of Marseille-1 variants

Marseille-1 variants (that we propose to name 20A.MAR1) have a backbone of 10 mutations when compared to the Nextrain clade 20A strain, including C241U (*nsp2* gene [unknown function], synonymous), G1181U (*nsp2* gene, amino acid substitution V126F), C1625U (*nsp2* gene, amino acid substitution L274F), C3037U (*nsp3* [phosphoesterase], synonymous), C14408U (*nsp12* [RNA-dependent RNA polymerase, RdRp], amino acid substitution P314L), G22894A (spike-coding [*S*] gene, synonymous), A23403G (*S* gene, D614G), C25886U (ORF3a [predicted phosphoesterase], amino acid substitution S165F), G28198U (ORF8 [unknown function], amino acid substitution C102F) (**Supplementary Figure 3**) and G28851U (nucleocapsid-coding [*N*] gene, amino acid substitution S1931I). By comparison with the Wuhan-Hu-1 strain, two additional mutations were noted, including A20268G (*nsp15*, synonymous) and C28833U (*N* gene, S187L; and ORF14, H34Y) (**Table 1**). Only amino acid substitutions V126F and L274F in the Nsp2 protein were found in the CoV-GLUE replacements database (http://cov-glue.cvr.gla.ac.uk/#/replacement) [11], in 123 and 308 GISAID genomes, respectively. The two additional mutations C22088U, corresponding to substitution L176F in the S protein and G5378A, corresponding to substitution G887S in the Nsp3 protein, were found in 612 and 9 GISAID genomes, respectively. In addition, we observed the successive occurrence of two additional mutations, C22088U (in Marseille-1A variants) then G5378A (in Marseille-1A1 variants) in 12 and 6, respectively, of the 38 full-length genomes obtained in Marseille. Moreover, 28 other mutations were found in between 1 and 6 of these Marseille-1 variant genomes, raising the total number of mutated nucleotide positions to 42 (**Table 1**). Interestingly, amino acid substitution C102F in ORF8 disrupts a disulfide bond close to the end of this 121 amino acid-long protein, which might alter its function, though as-yet undetermined.

By comparison with the original Wuhan-Hu-1 isolate, the Marseille-1 variant and its descendants (genotypes Marseille-1A and Marseille-1A1) differed by 12, 13 and 14 mutations, respectively. All three are classified in GISAID clade 20A and correspond to lineages B.1.5.12 and E.1 (an alias of B.1.5.12.1 lineage) in the Pangolin classification [12]. Their most recent identified ancestors are genomes from Senegal and Gambia that either harbored none of the 10 hallmark mutations, or C1625U, or C1625U associated to C25886U (**Figure 1**). This strongly suggests the evolution of Marseille-1 ancestors in these countries through the successive occurrence of these mutations.

### 3.5 Demographic and clinical features of patients infected with the Marseille-1 variant

We compared the characteristics of 336 patients infected between March and April 2020 with clade 20A strains and 81 patients infected with the Marseille-1 variant (**Table 2**). The patients infected with the Marseille-1 variant were more frequently to be male and younger than those infected with clade 20A strains from episode 1. Of the 417 patients, 56 were hospitalized. The hospitalization rate was lower in patients infected with the Marseille-1 variant. Ten patients died and five were transferred to intensive care unit, all of whom were infected with 20A variants. Clinical symptoms were available in 320 patients (Table 2). Patients infected with the Marseille-1 variant suffered less frequently from dyspnea and hypoxemia. In contrast, rhinitis, anosmia and ageusia were not significantly different between patients infected with either of the two variants. Overall, the Marseille-1 variant exhibited a milder phenotype [13].

**Table 2.**
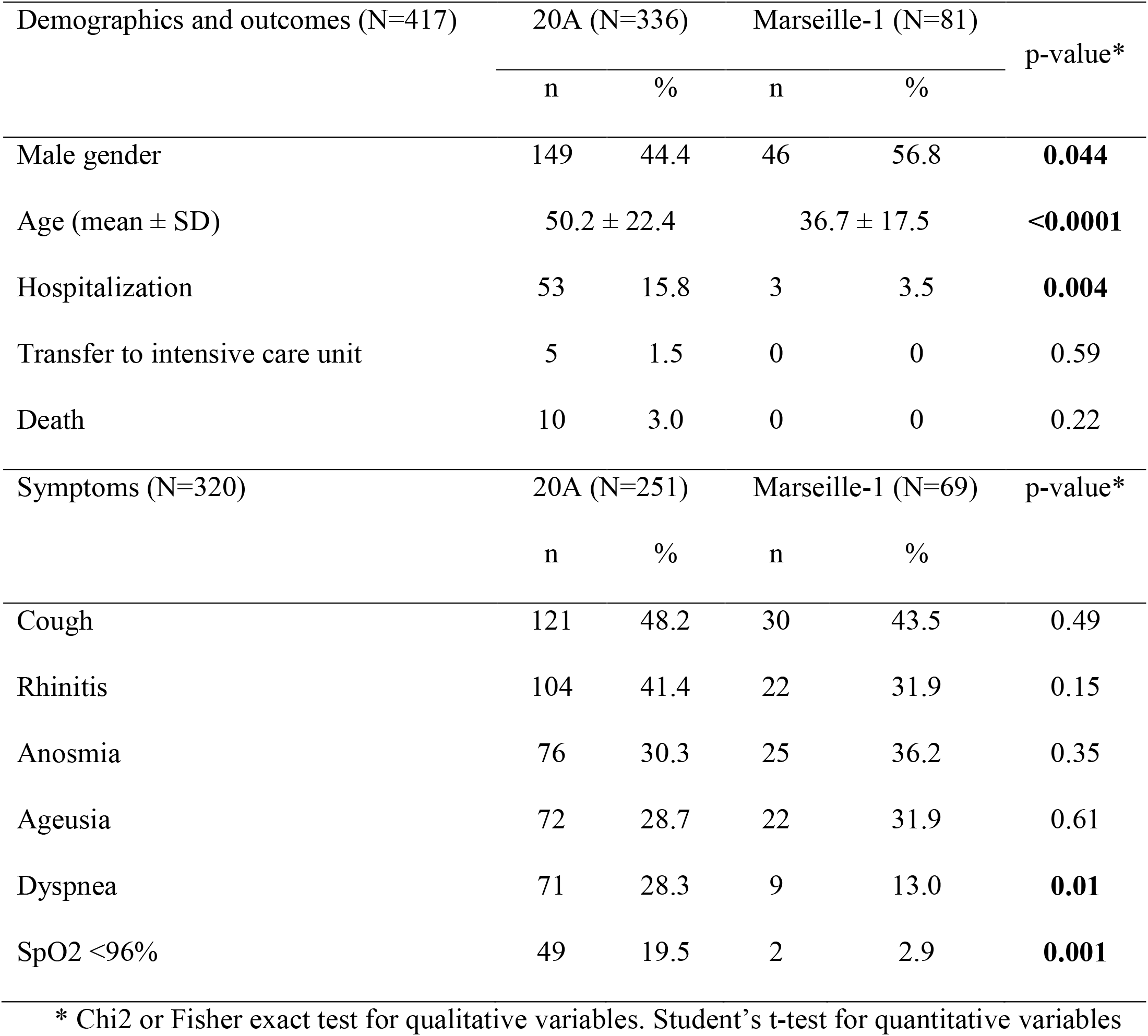
**Demographics, outcomes and clinical symptoms in patients infected with different SARS-CoV-2 variants**

| Demographics and outcomes (N=417) | 20A (N=336) |  | Marseille-1 (N=81) |  | p-value* |
| --- | --- | --- | --- | --- | --- |
|  | n | % | n | % |  |
| Male gender | 149 | 44.4 | 46 | 56.8 | **0.044** |
| Age (mean ± SD) | 50.2 ± 22.4 | | 36.7 ± 17.5 | | **<0.0001** |
| Hospitalization | 53 | 15.8 | 3 | 3.5 | **0.004** |
| Transfer to intensive care unit | 5 | 1.5 | 0 | 0 | 0.59 |
| Death | 10 | 3.0 | 0 | 0 | 0.22 |
| Symptoms (N=320) | 20A (N=251) |  | Marseille-1 (N=69) |  | p-value* |
|  | n | % | n | % |  |
| Cough | 121 | 48.2 | 30 | 43.5 | 0.49 |
| Rhinitis | 104 | 41.4 | 22 | 31.9 | 0.15 |
| Anosmia | 76 | 30.3 | 25 | 36.2 | 0.35 |
| Ageusia | 72 | 28.7 | 22 | 31.9 | 0.61 |
| Dyspnea | 71 | 28.3 | 9 | 13.0 | **0.01** |
| SpO2 <96% | 49 | 19.5 | 2 | 2.9 | **0.001** |

\* Chi2 or Fisher exact test for qualitative variables. Student's t-test for quantitative variables

## 4. Discussion

Here we demonstrate that two SARS-CoV-2 epidemic phenomena occurred in France. The first one, from March to May 2020, exhibited a usual evolution for a respiratory viral infection, and was similar to the one observed in China. In contrast, following an almost total disappearance of SARS-CoV-2 diagnoses, the second epidemic episode evolved in successive or overlapping waves. These waves resulted from the occurrence of 10 viral variants exhibiting substantial genetic diversity between each other. Among them, the Marseille-1 variant caused a short outbreak than ran from July to August 2020, and remained essentially restricted to the Marseille area in France. In addition, our study demonstrated that the Marseille-1 variant was present in sub-Saharan (Senegal, Gambia) and North Africa (Tunisia, Algeria, Morocco). This variant is most likely a descendant from clade 20A strains transferred to sub-Saharan Africa by French travelers during episode 1 [10, 14], prior to genetically evolving onsite and being later brought back to Maghreb and then to Marseille by travelers. The Marseille-1 variant was associated with a milder clinical outcome and a lower epidemic potential, and no associated death were observed. In addition, its epidemic potential was lower, and no case of re-infection with this variant was detected, contrasting with what we observed with other variants [15].

The evolution of the Marseille-1 variant as we understand it corresponds to a Darwinian model. Indeed, during episode 1, following the global spread of clade 20A strains, political responses have consisted in the interruption of travels and border closure. This resulted in the isolation of distinct populations and ecosystems for several months, in which viral variants may have evolved individually. However, we cannot rule out that the mutations observed in the Marseille-1 variant by comparison to the Chinese variant occurred in an animal population in sub-Saharan countries, as observed recently in farmed minks in Northern Europe [4]. Under these conditions, what generally occurs is speciation [16]. As a matter of fact, the existence of animal reservoirs, infected during the first episode, may explain the differences in epidemic curves observed among countries. However, the consequences of viral variant selection in massive animal groups, and subsequent human infections, remain to be determined, especially because the immunity acquired by patients during episode 1 may not be protective against a re-infection with another variant. [15]

Then, when international borders reopened and travels resumed, the reconnection of these isolated ecosystems where different variants had developed generated new outbreaks in areas that were the most exposed to incoming populations. This was in particular the case for Marseille where daily passenger air and boat traffic exchanges with Maghreb occur. Marseille is alas familiar with boat importation of epidemics from the South, notably plague and cholera [17]. Therefore, our results confirm that SARS-CoV-2 is able to genetically diversify rapidly, its variants to diffuse internationally with travelers and cause successive outbreaks, even in populations beforehand exposed to the original virus.

## Data Availability

All data used in the present manuscript are available upon request

## Author contributions

Conceived and designed the study: PC, DR. Designed and/or performed experiments: VTH, JD, MB, LB, VB, MBK. Analyzed patients’ data: PG, MM, JCL. Analyzed and interpreted data: PC, AL, PG, FF, IB, RS, MM, AP, HC, MM, HTD, JCL, SM, PEF, DR. Wrote the manuscript: PC, AL, PG, FF, MM, HC, MM, HTD, JCL, PEF, DR. All authors read and approved the final manuscript.

## Acknowledgments

We are grateful to Prs Philippe Brouqui and Philippe Parola for their involvement in the management of patients, and to Olivia Ardizzoni, Madeleine Carrera, Vera Esteves-Vieira, Laurence Thomas, Priscilla Jardot, Raphael Tola and Audrey Giraud-Gatineau for their technical help.

## References

[1] Colson P, Esteves-Vieira V, Giraud-Gatineau A, Zandotti C, Filosa V, Chaudet H, et al. Temporal and age distributions of SARS-CoV-2 and other coronaviruses, southeastern France. Int J Infect Dis 2020 Sep 23; 101:121–5. https://doi.org/10.1016/j.ijid.2020.09.1417

[2] Colson P, Lagier JC, Baudoin JP, Bou KJ, La SB, Raoult D. Ultrarapid diagnosis, microscope imaging, genome sequencing, and culture isolation of SARS-CoV-2. Eur J Clin Microbiol Infect Dis 2020 Aug; 39(8):1601–3. https://doi.org/10.1007/s10096-020-03869-w

[3] Fournier PE, Colson P, Levasseur A, Gautret P, Luciani L, Bedotto M, et al. Genome sequence analysis enabled to decipher the atypical evolution of COVID-19 epidemics in Marseille, France. bioRxiv 2020; PrePrint.

[4] Koeijer AA, Hagenaars TJ, Leuken JPGV, Swart AN, Boender GJ. Spatial transmission risk during the 2007-2010 Q fever epidemic in The Netherlands: Analysis of the farm-to-farm and farm-to-resident transmission. PLoS One 2020; 15(2):e0227491. https://doi.org/10.1371/journal.pone.0227491

[5] Lagier JC, Million M, Gautret P, Colson P, Cortaredona S, Giraud-Gatineau A, et al. Outcomes of 3,737 COVID-19 patients treated with hydroxychloroquine/azithromycin and other regimens in Marseille, France: A retrospective analysis. Travel Med Infect Dis 2020 Jun 25;101791. https://doi.org/10.1016/j.tmaid.2020.101791

[6] Levasseur A, Delerce J, Caputo A, Brechard L, Colson P, Lagier JC, et al. Genomic diversity and evolution of coronavirus (SARS-CoV-2) in France from 309 COVID-19- infected patients. bioRxiv 2020; Preprint. https://doi.org/10.1101/2020.09.04.282616

[7] Hadfield J, Megill C, Bell SM, Huddleston J, Potter B, Callender C, et al. Nextstrain: real-time tracking of pathogen evolution. Bioinformatics 2018; 34:4121–3. https://doi.org/10.1093/bioinformatics/bty407

[8] Lalaoui R, Bakour S, Raoult D, Verger P, Sokhna C, Devaux C, et al. What could explain the late emergence of COVID-19 in Africa? New Microbes New Infect 2020; 38:100760. https://doi.org/10.1016/j.nmni.2020.100760

[9] Dia N, Lakh NA, Diagne MM, Mbaye KD, Taieb F, Fall NM, et al. COVID-19 Outbreak, Senegal, 2020. Emerg Infect Dis 2020; 26:2772–4. https://doi.org/10.3201/eid2611.202615

[10] Wruck W, Adjaye J. Transmission of SARS-COV-2 from China to Europe and West Africa: a detailed phylogenetic analysis. bioRxiv 2020; Preprint. https://doi.org/10.1101/2020.10.02.323519

[11] Singer J, Gifford R, Cotten M, Robertson D. CoV-GLUE: A Web Application for Tracking SARS-CoV-2 Genomic Variation. Preprints 2020; 2020060225. https://doi.org/10.20944/preprints202006.0225.v1

[12] Rambaut A, Holmes EC, O’Toole A, Hill V, McCrone JT, Ruis C, et al. A dynamic nomenclature proposal for SARS-CoV-2 lineages to assist genomic epidemiology. Nat Microbiol 2020; 5:1403–7. https://doi.org/10.1038/s41564-020-0770-5

[13] Gautret P, Colson P, Lagier JC, Camoin-Jau L, Giraud-Gatineau A, Boudjema S, et al. Different pattern of the second outbreak of COVID-19 in Marseille, France. Int J Infect Dis 2020;102:17–9. https://doi.org/10.1016/j.ijid.2020.10.005

[14] Sun H, Dickens BL, Cook AR, Clapham HE. Importations of COVID-19 into African countries and risk of onward spread. BMC Infect Dis 2020; 20:598. https://doi.org/10.1186/s12879-020-05323-w

[15] Colson P, Finaud M, Levy N, Lagier JC, Raoult D. Evidence of SARS-CoV-2 re-infection with a different genotype. J Infect 2020; Epub. https://doi.org/10.1016/j.jinf.2020.11.011

[16] Darwin C. On the origin of species. London: John Murray, 1859.

[17] Barbieri R, Signoli M, Cheve D, Costedoat C, Tzortzis S, Aboudharam G, et al. Yersinia pestis: the Natural History of Plague. Clin Microbiol Rev 2020; 34. https://doi.org/10.1128/CMR.00044-19

[18] Kelley LA, Mezulis S, Yates CM, Wass MN, Sternberg MJ. The Phyre2 web portal for protein modeling, prediction and analysis. Nat Protoc 2015; 10:845–58. https://doi.org/10.1038/nprot.2015.053

