## Supplementary appendix for "Introduction into the Marseille geographical area of a mild SARS-CoV-2 variant originating from sub-Saharan Africa"

### TITLE PAGE

**Article type: Research article**

**Affiliations:** [1] IHU Méditerranée Infection, Marseille, France; [2] Aix-Marseille Univ, Microbes Evolution Phylogeny and Infections (MEPHI), Marseille, France; [3] Aix-Marseille Univ, Vecteurs - Infections Tropicales et Méditerranéennes (VITROME), Marseille, France; [4]Thai Binh University of Medicine and Pharmacy, Thai Binh, Vietnam, [5] Ecole Nationale Supérieure Vétérinaire d'Alger, Alger, Algeria; [6] Faculty of Sciences Ben M'sik, Hassan II University of Casablanca, Morocco ; [7] Institut de Recherche en Santé, de Surveillance Epidémiologique et de Formation (IRESSEF), Rufisque, Senegal; [8] French Armed Forces Center for Epidemiology and Public Health (CESPA), Marseille, France.

**Corresponding authors:** Didier Raoult, IHU - Méditerranée Infection, 19-21 boulevard Jean Moulin, 13005 Marseille, France. Tel.: +33 413 732 401, Fax: +33 413 732 402;

Pierre-Edouard Fournier, IHU - Méditerranée Infection, 19-21 boulevard Jean Moulin, 13005

### Supplementary Appendix

**Supplementary Figure 1. Weekly numbers of patients with COVID-19 diagnosed in the Méditerranée Infection institute and for whom the SARS-CoV-2 genotype was obtained by genome sequencing or variant-specific RT-PCR, and their geographical origin.** M1 indicates Marseille-1 variant; Other designates SARS-CoV-2 genotypes except Marseille-1.

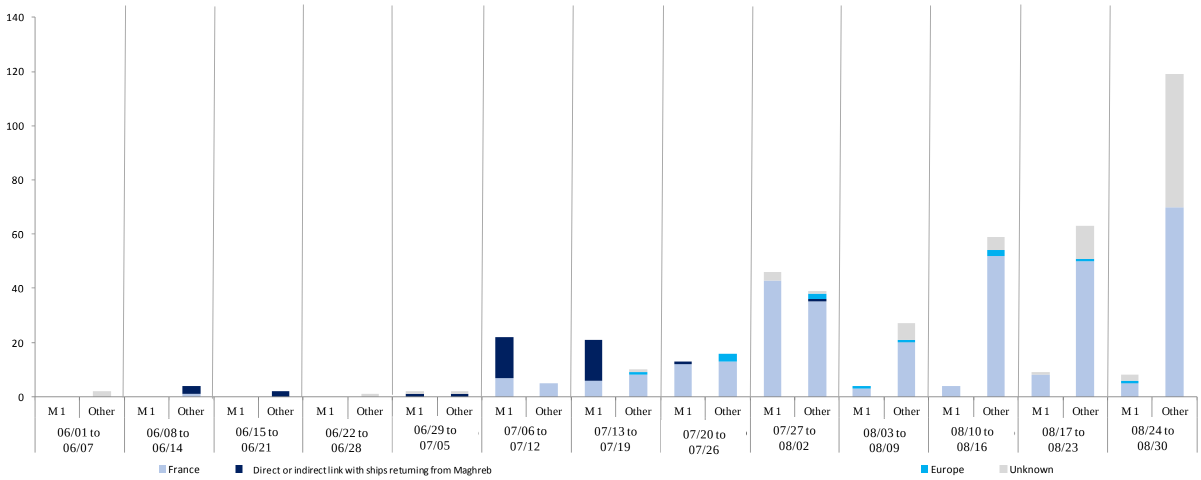

**Supplementary Figure 2. Genome sequence-based phylogenetic tree showing the evolution of Marseille-1 strains**

Genome sequences obtained in our institute were compared to those available in the GISAID database (https://www.gisaid.org/). Phylogenetic trees were reconstructed and visualized by using the Nextclade and iTOL (https://itol.embl.de/) web tools, respectively.

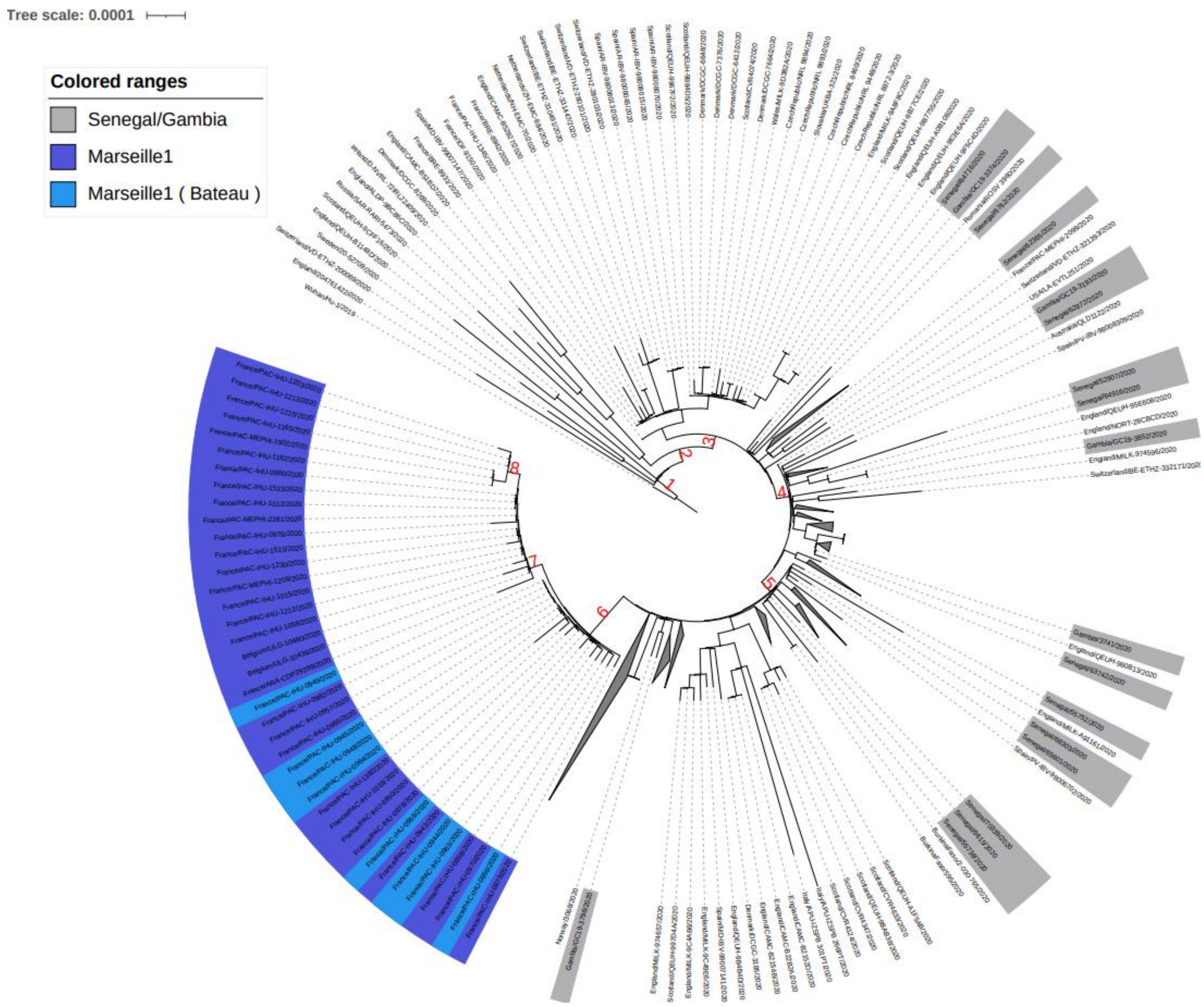

**Supplementary Figure 3. Amino acid alignment and structural prediction obtained using the Phyre2 web portal for protein modeling, prediction and analysis [18] for ORF8 gene product of the Marseille-1 variant.** This figure was obtained using the Phyre2 web portal (http://www.sbg.bio.ic.ac.uk/~phyre2/html/page.cgi?id=index).

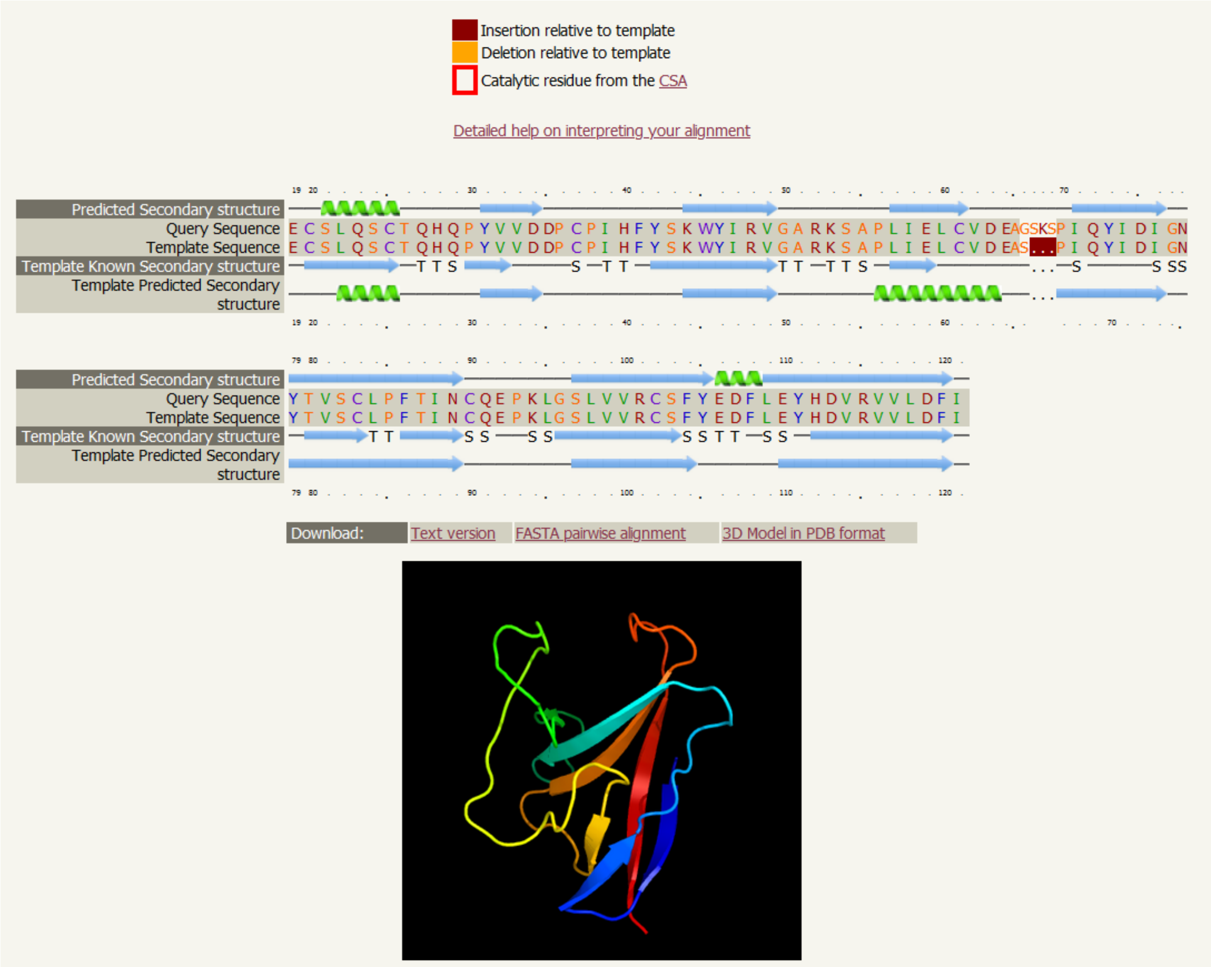

**Supplementary Table 1: Detail of commercial travels from Maghreb to France following the re-opening of French borders on June 1st, 2020.**

| Country | Date | Event | Source |
|---|---|---|---|
| **Algeria** | 25-05-2020 | One daily repatriation flight to Paris | https://www.dzairdaily.com/rapatriement-algerie-france-vol-quotidien-alger-paris-tarif-special-consulat-air-france/ |
| **Algeria** | 26-05-2020 | End of the repatriation of 56,000 French people (15,000 left) | https://www.leparisien.fr/societe/coronavirus-paris-augmente-les-vols-avec-le-maghreb-pour-rapatrier-15000-francais-26-05-2020-8324328.php |
| **Algeria** | 01-06-2020 | Reopening of the Lufthansa® and Swiss Air® lines | https://www.algerie-eco.com/2020/05/11/alger-francfort-et-alger-geneve-reouvertures-des-reservations-sur-lufthansa-et-swiss/ |
| **Algeria** | 01-06-2020 | Corsica Linea® maritime link: repatriation of 1000 people | https://www.diplomatie.gouv.fr/fr/le-ministere-et-son-reseau/actualites-du-ministere/informations-coronavirus-covid-19/coronavirus-declarations-et-communiques/article/mobilisation-renforcee-pour-faciliter-le-retour-en-france-des-ressortissants |
| **Algeria** | 05-06-2020 | Reopening of Air France flights from Algeria (Algiers, Oran, Annaba, Béjaïa, Constantine) to Paris. 4-5 flights/day for repatriation | https://www.diplomatie.gouv.fr/fr/le-ministere-et-son-reseau/actualites-du-ministere/informations-coronavirus-covid-19/coronavirus-declarations-et-communiques/article/mobilisation-renforcee-pour-faciliter-le-retour-en-france-des-ressortissants |
| **Algeria** | 07-06-2020 | Corsica Linea® maritime link: repatriation of 1000 people | https://www.diplomatie.gouv.fr/fr/le-ministere-et-son-reseau/actualites-du-ministere/informations-coronavirus-covid-19/coronavirus-declarations-et-communiques/article/mobilisation-renforcee-pour-faciliter-le-retour-en-france-des-ressortissants |
| **Algeria** | 09-06-2020 | Corsica Linea® maritime link: repatriation of 1000 people | https://www.diplomatie.gouv.fr/fr/le-ministere-et-son-reseau/actualites-du-ministere/informations-coronavirus-covid-19/coronavirus-declarations-et-communiques/article/mobilisation-renforcee-pour-faciliter-le-retour-en-france-des-ressortissants |
| **Algeria** | 27-06-2020 | Closure of air and sea passenger links | https://www.garda.com/crisis24/news-alerts/355311/algeria-president-announces-borders-to-remain-closed-until-end-of-covid-19-pandemic-update-20 |
| **Morocco** | 05-06-2020 | Reopening of La Méridionale® links between Nador and Marseille | https://www.diplomatie.gouv.fr/fr/le-ministere-et-son-reseau/actualites-du-ministere/informations-coronavirus-covid-19/coronavirus-declarations-et-communiques/article/mobilisation-renforcee-pour-faciliter-le-retour-en-france-des-ressortissants |
| **Morocco** | 05-06-2020 | Opening of 5 crossings until June 9th by company Balearia® for repatriation of 10,000 motorhome operators via Spain | https://www.diplomatie.gouv.fr/fr/le-ministere-et-son-reseau/actualites-du-ministere/informations-coronavirus-covid-19/coronavirus-declarations-et-communiques/article/mobilisation-renforcee-pour-faciliter-le-retour-en-france-des-ressortissants |
| **Morocco** | 08-06-2020 | Reinforced program of 60 flights between Casablanca or Marrakech and Paris until June 25th for French people repatriation (total 10,000) | https://www.diplomatie.gouv.fr/fr/le-ministere-et-son-reseau/actualites-du-ministere/informations-coronavirus-covid-19/coronavirus-declarations-et-communiques/article/mobilisation-renforcee-pour-faciliter-le-retour-en-france-des-ressortissants |
| **Morocco** | 09-06-2020 | Opening of company GNV® passenger link between Tangier and Sète | https://www.diplomatie.gouv.fr/fr/le-ministere-et-son-reseau/actualites-du-ministere/informations-coronavirus-covid-19/coronavirus-declarations-et-communiques/article/mobilisation-renforcee-pour-faciliter-le-retour-en-france-des-ressortissants |
| **Morocco** | 06-09-2020 | Opening of the borders | https://www.quotidiendutourisme.com/actualite/destinations/le-maroc-rouvre-ses-frontieres-aux-touristes-francais-605318.php |
| **Tunisia** | 04-06-2020 | Corsica Linea® repatriation link 1000 pax | https://www.diplomatie.gouv.fr/fr/le-ministere-et-son-reseau/actualites-du-ministere/informations-coronavirus-covid-19/coronavirus-declarations-et-communiques/article/mobilisation-renforcee-pour-faciliter-le-retour-en-france-des-ressortissants |
| **Tunisia** | 05-06-2020 | 2 weekly repatriation flights | https://www.diplomatie.gouv.fr/fr/le-ministere-et-son-reseau/actualites-du-ministere/informations-coronavirus-covid-19/coronavirus-declarations-et-communiques/article/mobilisation-renforcee-pour-faciliter-le-retour-en-france-des-ressortissants |
| **Tunisia** | 11-06-2020 | Corsica Linea® repatriation link 1000 pax | https://www.diplomatie.gouv.fr/fr/le-ministere-et-son-reseau/actualites-du-ministere/informations-coronavirus-covid-19/coronavirus-declarations-et-communiques/article/mobilisation-renforcee-pour-faciliter-le-retour-en-france-des-ressortissants |
| **Tunisia** | 27-06-2020 | Reopening of borders | https://www.facebook.com/santetunisie.rns.tn/posts/3144668892238860?__cft__[0]=AZVaQeBvNr1QplZ5hPtt91WzVLWjjz5Vlebruy7gCYLTAHQBl6zSdm4jKkg1iZ743TBO7fgU8mFkeh0OULz1cMAfYRp49neCNeiyJ77GCt_pCABhK2OkKTOsz8onjdVcZPksW8VJ7pSt-xj0pF2ZdGUcuZp3-dcm_eGlmADU69qSBWU4vK5C4HlXJoGeoTPFvpe1k1CefIGThadtllqMfzzKedG2KIasasdUzGTzg0AzkA&__tn__=%2CP-R |
| **Tunisia** | 29-06-2020 | Resumption of ferry links (around 700 passengers per rotation) | https://www.webmanagercenter.com/2020/06/29/452895/transport-maritime-reprise-du-trafic-maritime-au-port-de-zarzis/ |
